# Caffeine Consumption and Schizophrenia: A Systematic Review and Meta-analysis of Cognitive, Symptomatic, and Functional Outcomes

**DOI:** 10.1101/2025.06.16.25328846

**Authors:** Zhen Zhou Benjamin Wong, Qi Xu Soh, Aaron W.K. Mau, Wei Hong Xia, Qi Rui Soh

## Abstract

**Background:** Schizophrenia is a complex mental disorder characterized by cognitive deficits, persistent symptoms, and functional impairment. Caffeine, a commonly consumed psychoactive substance, has plausible effects on cognition and mood. However, its impact on individuals with schizophrenia remains unclear. This review evaluates caffeine’s impact on cognition, symptomatology, and functional outcomes in schizophrenia.

**Method:** A systematic literature search was conducted for articles published up to 30 December 2024 across PubMed, Cochrane Library, PsycINFO, Embase, Emcare, and Medline. We included English-language studies in adults with schizophrenia that compared different caffeine intake levels and reported outcomes on cognition, symptoms, or functioning. Cohort, cross-sectional, and clinical trial designs were included. Data were synthesized using a random-effects model with Hedges’ g for effect size and I² statistics for heterogeneity.

**Results:** Of 252 articles screened, eleven studies (n=1,406) met inclusion criteria. Findings were mixed. Some studies reported improvements in cognitive performance and working memory, while others observed increases in positive symptoms or inconsistent associations with overall symptom management. Meta-analyses revealed a non-significant increase/decrease in overall symptom severity (measured with Brief Psychiatric Rating Scale (BPRS) and Nurses’ Observation Scale for Inpatient Evaluation (NOSIE). Physiologically, caffeine was found to reduce cerebral blood flow, with no statistically significant effects on blood pressure or pulse.

**Conclusion:** Caffeine may have mixed effects on schizophrenia, with potential positive effects on cognitive and negative symptoms while possibly worsening positive symptoms. Functional and physiological impacts are unclear, warranting further research to guide clinical recommendations.

## 1. Introduction

Schizophrenia is a chronic and severe mental health disorder that manifests through a broad spectrum of cognitive, emotional, and behavioural impairments (Keefe and Harvey, 2012, Elvevag and Goldberg, 2000, Keefe and Fenton, 2007). Globally, approximately 29 million individuals live with schizophrenia, placing significant burdens on patients, their families, and healthcare systems (Chan, 2011). Notably, cognitive deficits are a core feature of the disorder, often persisting despite other symptom remission. These deficits significantly impact functional outcomes such as employment, social interactions, and independent living (Rihmer et al., 2015, Millan et al., 2012). Thus, targeted interventions are urgently needed to enhance quality of life and improve long-term prognosis.

Caffeine is the most widely consumed psychoactive substance in the modern world (Temple et al., 2017, Weinberg and Bealer, 2004). Millions worldwide rely on caffeine for its effects on alertness and mood. Although its consumption spans all ages, genders, and walks of life, those with psychiatric illnesses—such as schizophrenia— tend to consume it more (Marthoenis and Jannah, 2024). Caffeine, which can enhance cognitive performance and modulate mood, was thus hypothesized to influence symptoms and functioning in schizophrenia, yielding either therapeutic or deleterious effects depending on the context. The relationship is complex, with studies suggesting that caffeine may affect neurotransmitter systems implicated in the disorder, such as dopamine and adenosine pathways (Reddy et al., 2024). On one hand, caffeine has stimulant properties that act against cognitive fatigue and improve concentration (Cappelletti et al., 2015). Conversely, there are concerns that caffeine could exacerbate psychotic symptoms, anxiety, or sleep difficulties, complicating its overall effect (Temple et al., 2017, Radwan et al., 2022). Despite these historical insights, the specific effects of caffeine on cognitive symptoms, symptom management, and functional outcomes in schizophrenia remain underexplored and poorly understood.

Previous studies have primarily investigated caffeine’s effects on cognitive performance, symptomatology, and functional outcomes, mainly in general populations and those with other psychiatric conditions. Among non-psychiatric subjects, caffeine consistently enhances cognitive domains such as attention, reaction time, and working memory (Kennedy and Wightman, 2022). This was attributed to its antagonism of adenosine receptors and subsequent modulation of dopaminergic activity (Collins et al., 2010). Clinically, there has been research exploring caffeine as an adjunct therapy, noting benefits in managing depressive symptoms and reducing cognitive fatigability (Bao et al., 2022, Lopresti, 2019).

However, evidence about caffeine’s effects in schizophrenia is both limited and inconsistent. These conflicting findings support the need for investigation into a threshold between beneficial and harmful levels of caffeine intake in this population. Vitally, much of the existing research is nonspecific, with few studies focusing exclusively on individuals with schizophrenia and no systematic reviews comparing different levels of caffeine consumption on cognitive symptoms, overall symptom management, and functional outcomes in this specific patient population.

These gaps underscore the need for a comprehensive synthesis of the available evidence to clarify the role of caffeine, inform future research, and guide clinical practice. With caffeine use so pervasive among those with schizophrenia, it is imperative that we fully explore and understand its effects. Given its established cognitive-enhancing properties, it may offer a low-cost, widely available intervention to help address these cognitive deficits. However, without systematic study, its risks, such as exacerbation of psychotic symptoms or sleep disturbances, remain poorly defined. Therefore, this systematic review and meta-analysis seems to evaluate the impact of caffeine consumption compared to no or lower caffeine intake on cognitive symptoms, overall symptom management, and functional outcomes in patients with schizophrenia.

### 1.1 Research Question

In individuals with schizophrenia, how does caffeine consumption compared to no or lower caffeine intake affect cognitive symptoms, symptom management, and overall functioning?

## 2. Methods

### 2.1 Study Design

The study was registered with the PROSPERO international database of prospectively registered systematic reviews (CRD42025628484) (Booth et al., 2012). Preferred Reporting Items for Systematic Review and Meta-Analysis (PRISMA) statement recommendations were followed for the background, search strategy, methods, results, discussion, and conclusions (Page et al., 2021).

### 2.2 Information Sources and Study Selection

A systematic literature search was conducted for articles published up to 30^th^ December 2024. The index databases used were PubMed, Medline, Cochrane Library, PsycINFO, Embase, and Emcare. Topic keywords were used to generate search strings. The keywords utilized were “Schizophrenia,” “Schizophrenic,” “Caffeine,” “coffee,” “symptoms”. Relevant articles and their full text were collected for further assessment. We have created a PRISMA chart that depicts the flow of inclusion and exclusion of articles with reasoning [**Figure 1**].

**Figure 1:**
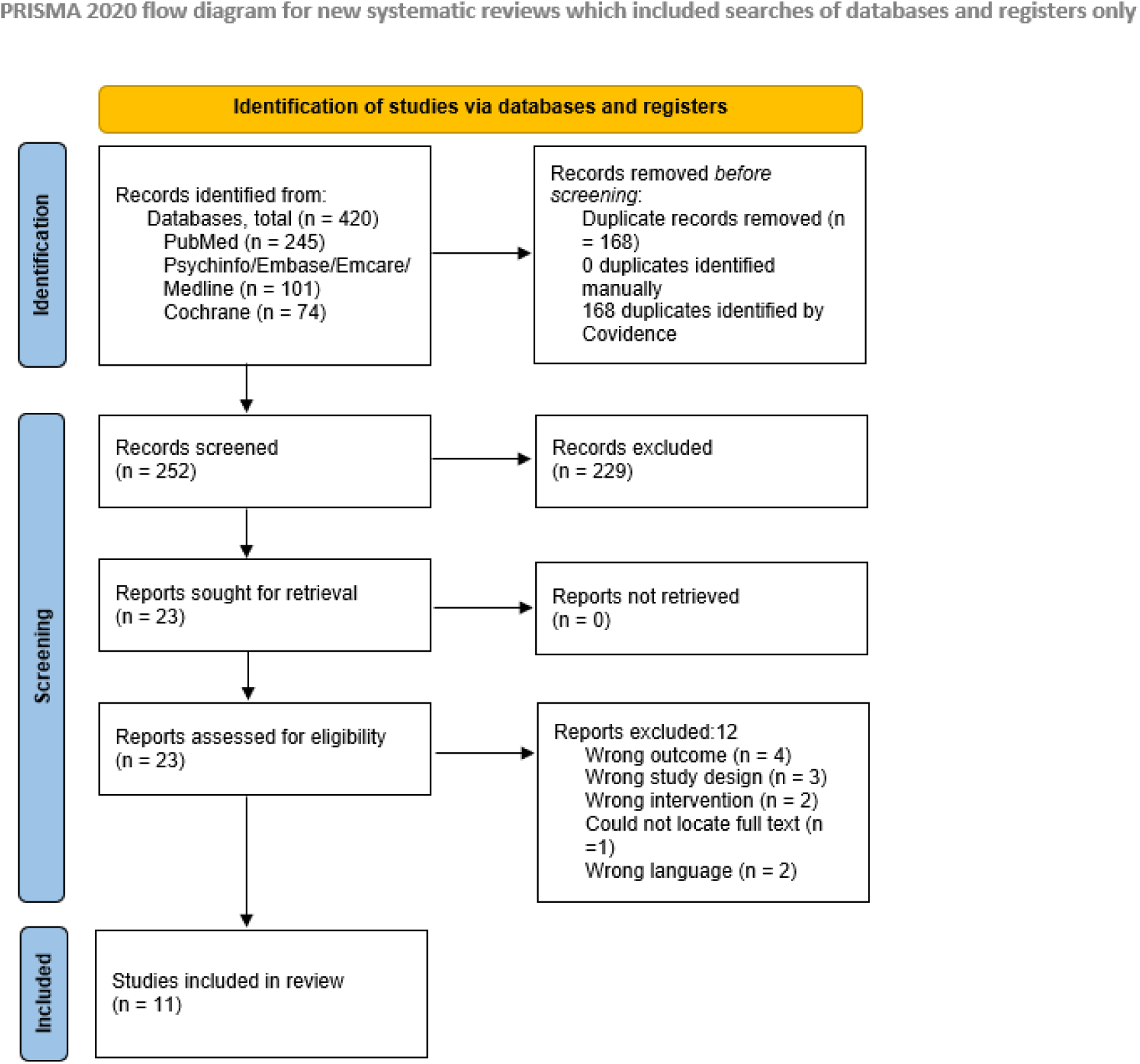
PRISMA flowchart showing search strategy and study selection process.

### 2.3 Inclusion and Exclusion Criteria

Studies were included in the review if they fulfilled the following criteria: (i) studies written in English and available in full text; (ii) studies involving individuals aged 18 years or older diagnosed with schizophrenia; (iii) studies evaluating caffeine consumption as the primary variable; (iv) comparisons of different levels of caffeine intake (e.g., high vs. low or none); (v) studies reporting on at least one of the following: cognitive symptoms, overall symptom management, and functional outcomes; (vi) be prospective or retrospective cohort studies, cross-sectional studies, or clinical trials.

Non-journal articles, conference abstracts, non-full text articles, systematic reviews, meta-analyses, or other secondary research papers were excluded. Animal studies or in vitro research were also excluded.

### 2.4 Review of Methodological Quality

The Joanna Briggs Institute (JBI) critical appraisal checklist tool for cohort was used to appraise the observational studies (Aromataris E, 2024). The tool contains several questions that can be answered as either Yes, No, Unclear, or NA. The overall quality of the study is determined by the culmination of these answers. JBI checklist for cross-sectional studies was also used to appraise cross-sectional studies(Moola S, 2020). Clinical trial studies were appraised using the Risk of Bias 2 (RoB 2) tool. The tool focused on aspects such as randomization, deviation from intended interventions, missing data, and measurement of outcomes(Sterne et al., 2019). Each study was independently assessed by two reviewers and in case of any discrepancies, they were resolved through a discussion and consensus.

### 2.5 Data Extraction

Data from the eligible articles were extracted into a predefined Excel sheet. The data extracted included the first author’s name and year of publication, study design, study region, sample size (% male), mean age (mean ± SD) years, caffeine exposure, comparison group, outcome measures, and results.

### 2.6 Statistical Analysis

Revman 9.2.1 was used to perform the meta-analysis. The outcomes of Brief Psychiatric Rating Scale(BPRS) and Nurses’ Observation Scale for Inpatient Evaluation (NOSIE) scores were analysed using a forest plot to obtain the Hedges’ g, 95% Confidence Interval (95% CI) and the p-values. Dichotomous outcomes (blood pressure and pulse) had their mean and standard deviation pooled for analysis too. For all analyses, if mean and standard deviation were not available, the median was considered equivalent to the mean, the interquartile range was converted to standard deviation by dividing by 1.35 and the range was transformed to standard deviation by dividing by 4, per the Cochrane handbook (Higgins et al., 2011). Statistical heterogeneity was assessed using I^2 statistics and Cochran Q test values, with values of 25%, 50%, and 75% indicating low, moderate and high heterogeneity respectively. Publication bias was assessed using funnel plots. A random effects model(hartung-knapp-sidik-jonkman method) was used and a p-value of 0.05 was employed as the significance threshold.

### 2.7 Ethical Considerations

As this study is based on data from already published studies, no ethical approval was required. All included studies were assumed to have obtained ethical approval.

## 3. Results

### 3.1 Literature Search

The initial search yielded a total of 420 articles: 245 from PubMed, 74 from Cochrane Library, 101 from Psychinfo/Embase/Emcare/Medline. 168 duplicates were removed, and the remaining 252 articles were subjected to a title and abstract screening. 229 articles were then excluded for topic irrelevance and 23 were subjected to a full-text review. Eleven studies were found eligible according to the eligibility criteria and included in this review [**Figure 1**].

### 3.2 Baseline Characteristics

This paper included eleven studies, comprising four cohort studies (Han Almis et al., 2023, Koczapski et al., 1989, Szoke et al., 2023, Núñez et al., 2015), four clinical studies (N. Bissonnette et al., 2022, Lucas et al., 1990, Mathew et al., 1986, Mayo et al., 1993), and three cross-sectional studies (Lagreula et al., 2023, Thompson et al., 2014, Topyurek et al., 2020). The total sample comprised 1,406 participants, including both individuals with schizophrenia and control subjects. Males constituted most of the study populations. The mean age of participants was 37.9 ± 10.7 years, with samples ranging from 13 (Lucas et al., 1990) to 804 (Szoke et al., 2023). The researchers involved different geographical areas, including Canada, Turkey, Belgium, the US, the UK, Spain, France, and Australia. The outcomes presented were SAPS, SANS, PANSS, BNSS, PSYRATS, BPRS, NOSIE, GAF, CAS, and CBS [Supplementary Table 1].

### 3.3 Quality Assessment and Publication Bias

The quality assessment results demonstrated that the majority of included studies were of good quality across the evaluated study designs. For cohort studies, four studies were assessed using the JBI tool checklist. All studies met all 11 criteria [**Supplementary Table 2**]. For cross-sectional studies, three studies were assessed using the JBI tool checklist. Two studies had an overall good quality and one study was deemed fair [**Supplementary Table 3**]. Four randomised controlled trials were assessed using the RoB 2 tool. Three studies had an overall “low risk of bias” and one study had an overall “some concerns” [**Figure 2**].

**Figure 2:**
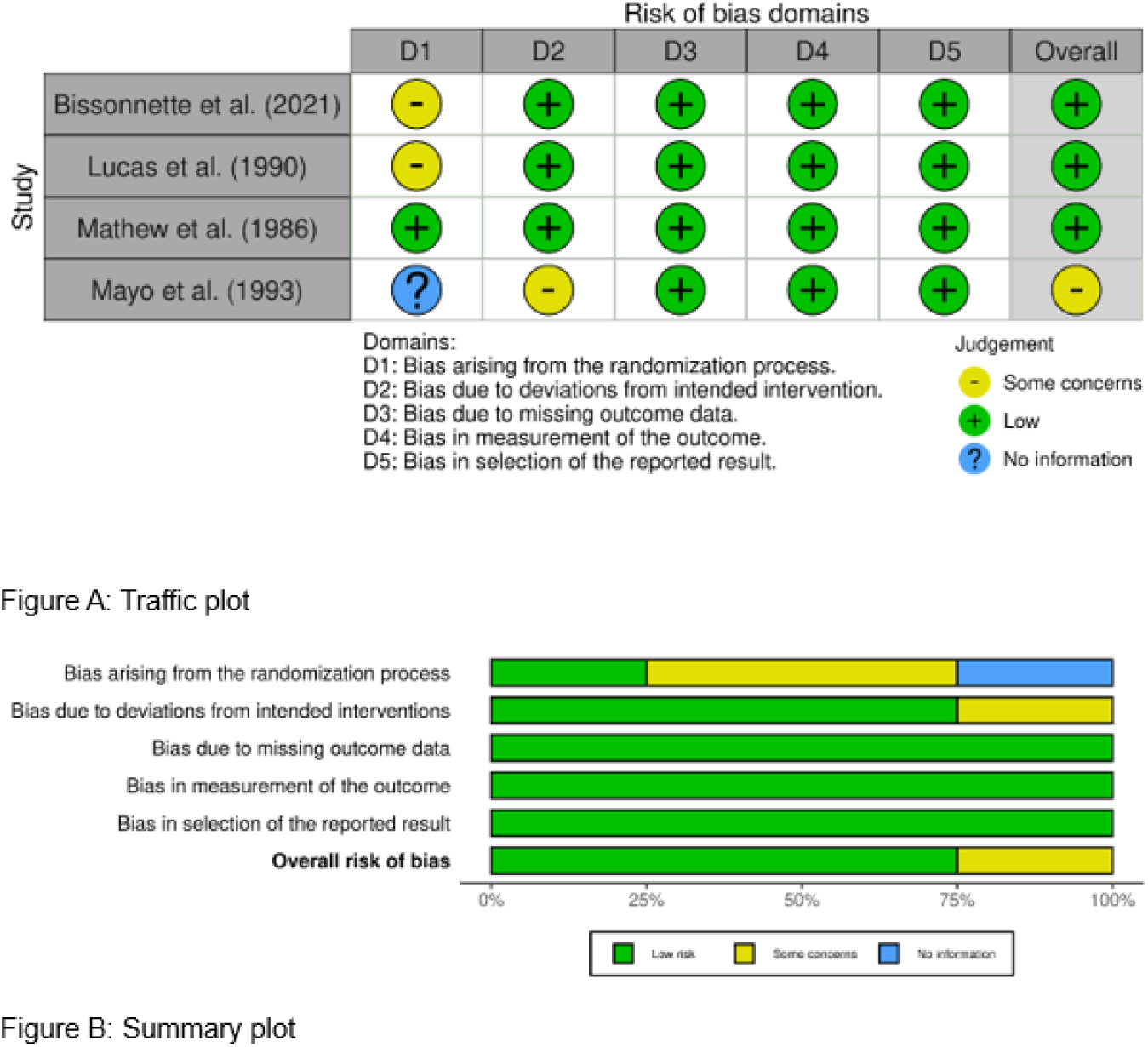
Bias assessment for RCT studies using RoB 2 tool.

Overall, these studies were found to be of high quality, with a low risk of bias. The funnel plots were considered sufficiently symmetrical to warrant inclusion in the meta-analysis.

### 3.4 Results of included studies

#### 3.4.1 Cognitive Symptoms

The impact of caffeine consumption on cognitive symptoms among individuals with schizophrenia was explored in three of the included studies, with mixed findings. Mathew et al. examined changes in patients’ cognitive status following caffeine administration and reported no observable changes in mental status when compared to placebo, suggesting a neutral effect of caffeine on overall cognitive functioning in this early study (Mathew et al., 1986). In contrast, Topyurek et al. found that caffeine consumption may influence executive function, as demonstrated in performance on the Groton Maze Learning Test. Specifically, there was a statistically significant difference where moderate caffeine users made significantly fewer errors compared to high caffeine users, suggesting that higher caffeine intake might be associated with diminished executive function (F(1, 25) = 7.5, p < 0.05, η² = 0.2) (Topyurek et al., 2020).

Lastly, Núñez et al. provided nuanced insights into the impact of caffeine on cognition in schizophrenia. They found that caffeine intake was positively associated with cognitive performance in specific domains, including cognitive/motor speed, working/short-term memory, and visual memory — with the notable exception of verbal memory (Núñez et al., 2015). These beneficial effects were observed exclusively in male patients, particularly among younger males, while no significant associations were found in female patients or in healthy controls of either sex.

Overall, the evidence suggests that caffeine’s effects on cognitive symptoms in schizophrenia may vary depending on dosage, habitual use, and patient characteristics such as sex and age.

#### 3.4.2 Symptom Management

Several studies have explored the relationship between caffeine consumption and the symptom profile of individuals with schizophrenia, considering both positive and negative dimensions.

Negative symptoms of schizophrenia were found to be associated with caffeine intake in two studies. Szoke et al. found a small but statistically significant negative correlation between current caffeine consumption and the negative dimension of the Positive and Negative Syndrome Scale (PANSS), suggesting that higher caffeine intake is linked to lower negative symptom scores (correlation coefficient = -0.10) (Szoke et al., 2023). Similarly, Topyurek et al. observed that high caffeine users had significantly fewer negative symptoms compared to moderate users (F(1, 25) = 7.7, p < 0.05, η² = 0.2), supporting the idea that caffeine may alleviate negative symptoms (Topyurek et al., 2020). Topyurek et al. also reported on the correlation between caffeine use and positive symptoms, with high caffeine users displaying significantly more positive symptoms than moderate users (F(1, 25) = 5.3, p < 0.05, η² = 0.2). This suggests a potential trade-off in which improvements in negative symptoms could come at the cost of exacerbated positive symptoms.

In contrast, Almis et al. did not observe any significant correlations between daily caffeine consumption and either positive or negative symptom scores, as measured by the Scale for the Assessment of Positive Symptoms (SAPS) and the Scale for the Assessment of Negative Symptoms (SANS) (Han Almis et al., 2023). Interestingly, they noted that female patients had significantly higher SAPS-bizarre behaviour scores than male patients (p = 0.02), indicating possible gender-related symptom expression, although unrelated directly to caffeine intake.

Further complicating the picture, exploratory neurophysiological investigations by Bissonnette et al. suggested that individuals with pronounced negative symptoms might exhibit stronger caffeine-induced effects on resting theta brainwave power, while those with auditory hallucinations demonstrated a reduced caffeine-related effect on beta and alpha_2_ brainwave power. Although these neurophysiological effects were not directly correlated with symptom scores, they hint at complex underlying mechanisms (N. Bissonnette et al., 2022).

Overall, these findings indicate a complex and inconsistent relationship between caffeine intake and symptoms of schizophrenia. While there is some evidence that caffeine might be beneficial for alleviating negative symptoms, it may concurrently exacerbate positive symptoms.

##### BPRS score

A meta-analysis was conducted to quantitatively assess the effect of caffeine on overall psychiatric symptom severity, as measured by the Brief Psychiatric Rating Scale (BPRS). Three studies were included in this analysis (Koczapski et al., 1989, Lucas et al., 1990, Mayo et al., 1993), comparing individuals with schizophrenia who consumed caffeine (experimental group) to those who did not (control group). The pooled standardized mean difference (SMD) was Hedges’ g = 0.92; 95% CI [-1.33, 3.17]; p < 0.001 [Figures 3 and 4], indicating a non-significant trend toward higher overall symptom severity in caffeine users compared to non-users. Notably, substantial heterogeneity was observed across studies (I² = 93%).

**Figure 3:**
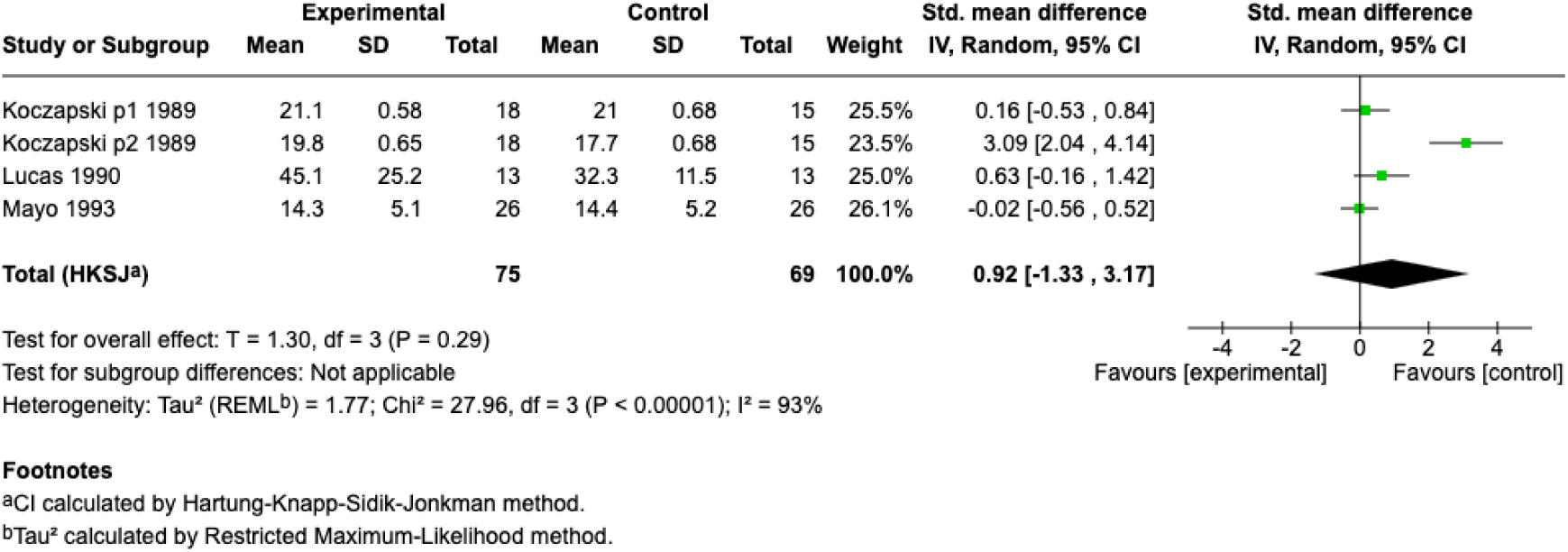
Forest plot of BRPS comparing experimental vs control groups in three studies.

**Figure 4:**
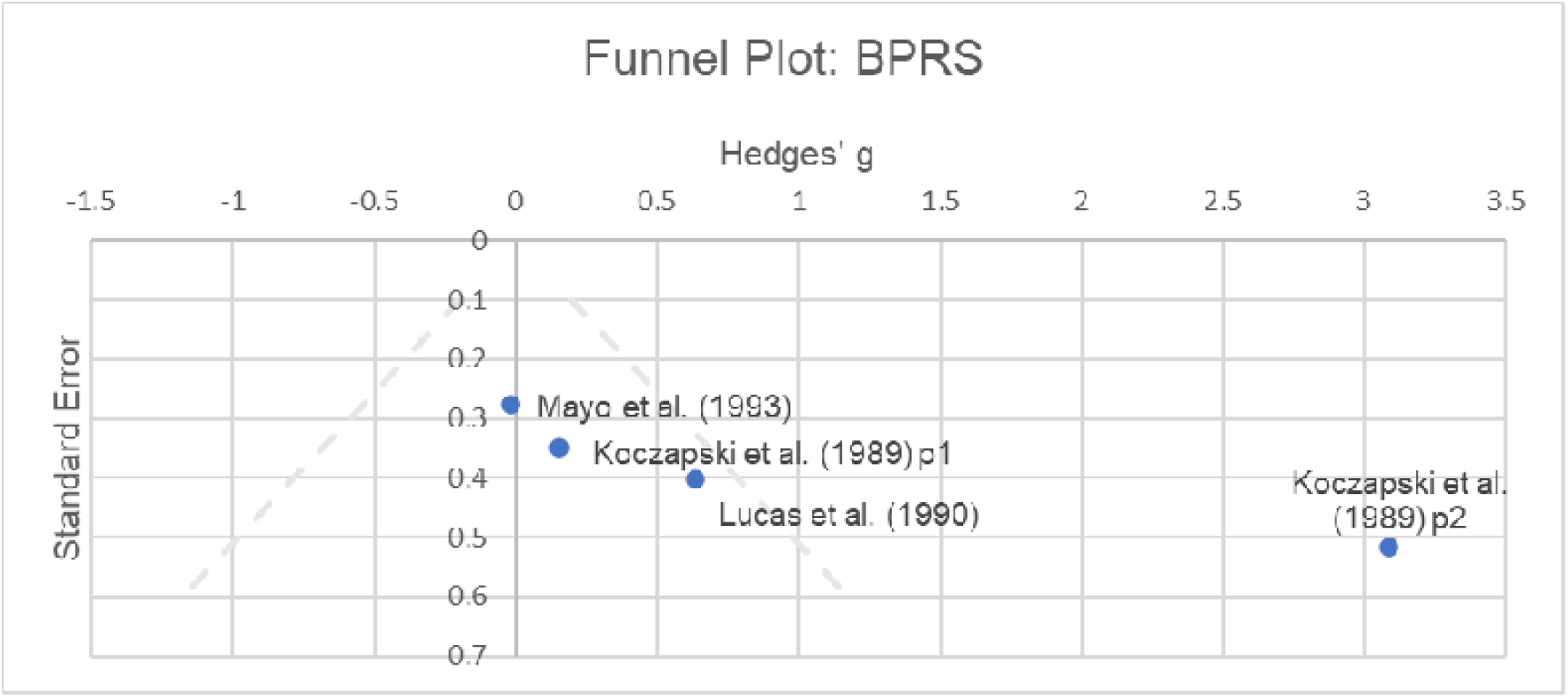
Funnel plot of Hedges’ g vs Standard error in BRPS of publications.

##### NOSIE score

Furthermore, a meta-analysis was performed on the Nurses’ Observation Scale for Inpatient Evaluation (NOSIE) to assess the impact of caffeine consumption on observed behavioural and psychiatric symptoms in individuals with schizophrenia. This analysis included two studies (Koczapski et al., 1989, Mayo et al., 1993). The pooled standardized mean difference (SMD) was Hedges’ g = -0.25; 95% CI [-1.52, 1.01]; p = 0.09 [Figures 5 and 6], indicating a small, non-significant trend toward lower symptom severity in caffeine users compared to non-users. The analysis showed moderate heterogeneity (I² = 58%). These findings suggest that, based on the limited available data, caffeine consumption does not have a consistent or significant effect on behaviour and observable symptoms among individuals with schizophrenia, although moderate variability between studies exists.

**Figure 5:**
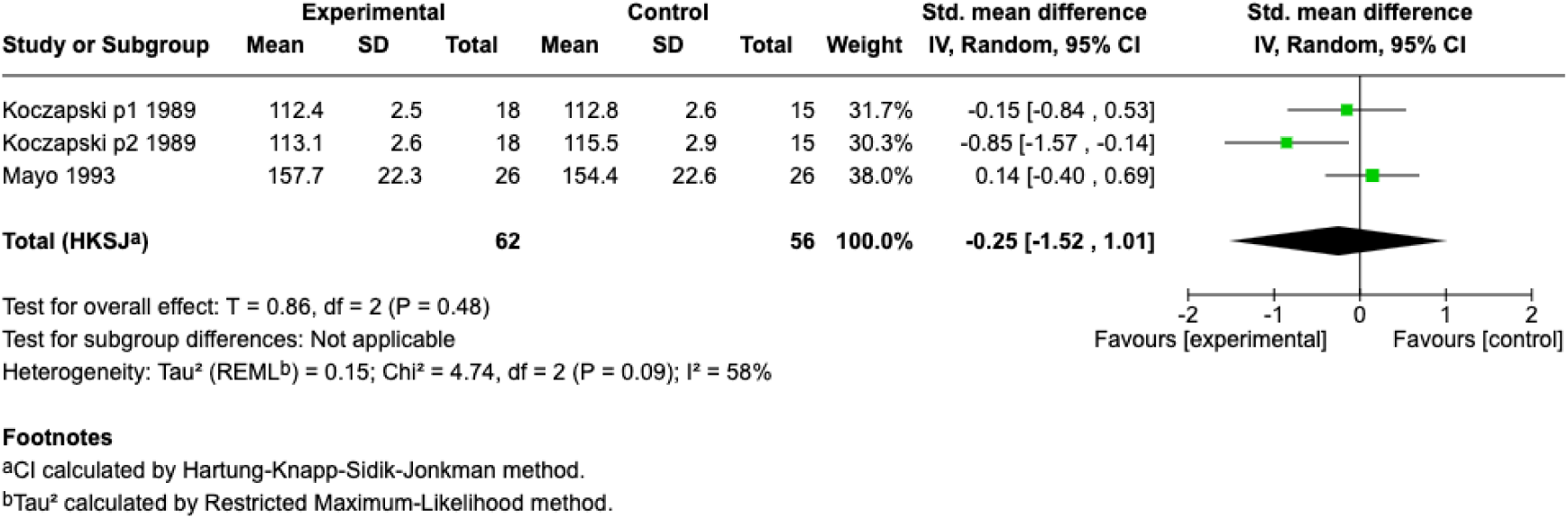
Forest plot of NOSIE score comparing experimental vs control groups in two studies.

**Figure 6:**
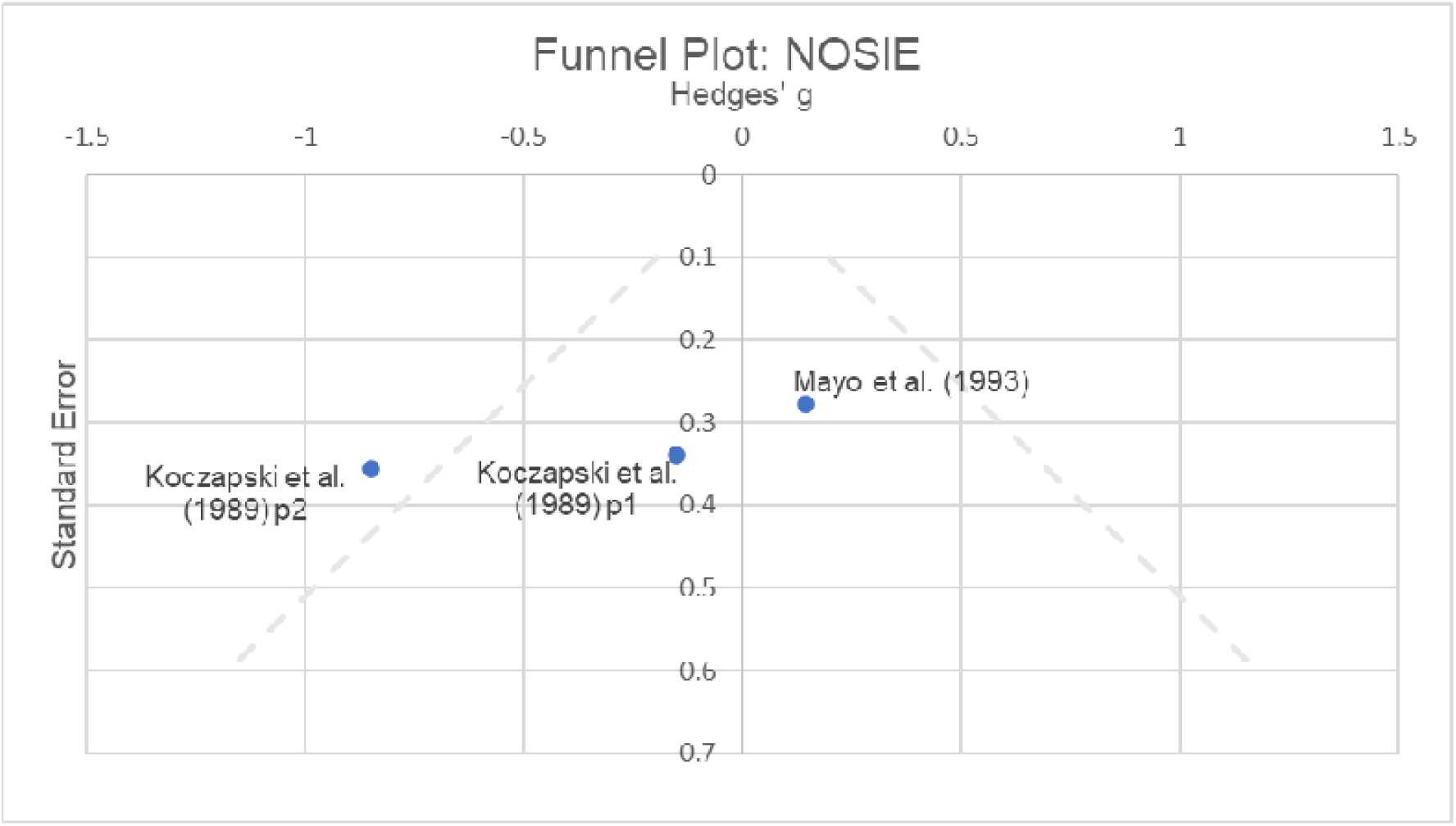
Funnel plot Hedges’ g vs Standard error in NOSIE score of publications.

#### 3.4.3 Functional Outcomes

Although evidence on caffeine’s impact on functional outcomes in schizophrenia is limited, some studies reported noteworthy findings. Lagreula et al. observed that restricted caffeine access within a hospital setting was significantly associated with better functional outcomes at discharge, as measured by the Global Assessment of Functioning (GAF) scale. After adjusting for potential confounding variables, patients in the decaffeinated group had significantly higher GAF scores upon discharge (adjusted odds ratio [aOR] = 2.86; 95% CI [1.77–4.62]), suggesting improved overall functioning compared to those with unrestricted caffeine access. Interestingly, despite having lower GAF scores upon admission (aOR = 0.42; 95% CI [0.26– 0.67])—indicating worse baseline functioning—patients in the decaffeinated group demonstrated greater improvement throughout their hospital stay. Additionally, a shorter length of hospitalization was associated with periods of restricted caffeine access (aOR = 0.68; 95% CI [0.47–0.99]), suggesting that reduced caffeine availability may contribute to more efficient clinical recovery and earlier discharge(Lagreula et al., 2023).

On the contrary, Szoke et al. reported that higher caffeine consumption was associated with lower negative and total PANSS scores, which in turn correlated with better clinical functioning. Specifically, caffeine consumption was associated with less severe clinical symptoms and improved functional status, as reflected by better Clinical Global Impression (CGI) and GAF scores in adjusted analyses (Szoke et al., 2023).

Findings regarding caffeine’s impact on hospitalization outcomes remained neutral in other studies. Mayo et al. found no significant association between caffeine consumption and length of hospital stay, highlighting the variability in research conclusions on this topic (Mayo et al., 1993).

#### 3.4.4 Physiological outcomes

Although only a few studies have examined the physiological effects of caffeine in individuals with schizophrenia, one notable investigation by Mathew et al. found that caffeine consumption significantly reduced cerebral blood flow (CBF) (Mathew et al., 1986). This reduction occurred across both hemispheres and most brain regions—a phenomenon not observed following placebo administration, suggesting a possible influence of caffeine on cerebral hemodynamics.

In addition to assessing cerebral blood flow, three meta-analyses were conducted to evaluate the effects of caffeine on systolic blood pressure, diastolic blood pressure, and pulse rate. For each outcome, data were pooled from two studies (Lucas et al., 1990, Mathew et al., 1986). The meta-analysis on systolic blood pressure indicated a small, non-significant increase among caffeine users compared to controls (Hedges’ g = 0.40; 95% CI [-0.48, 1.27]; p = 0.81), with no heterogeneity (I² = 0%). Similarly, diastolic blood pressure showed a non-significant increase in the caffeine group (Hedges’ g = 0.36; 95% CI [-1.53, 2.26]; p = 0.60), with no heterogeneity (I² = 0%). In terms of pulse rate, the pooled analysis showed a moderate but non-significant increase in the caffeine group (Hedges’ g = 0.74; 95% CI [-0.82, 2.31]; p = 0.68), also with no heterogeneity (I² = 0%). Forest plots illustrating these analyses are presented in Figure 7C-E

**Figure 7A:**
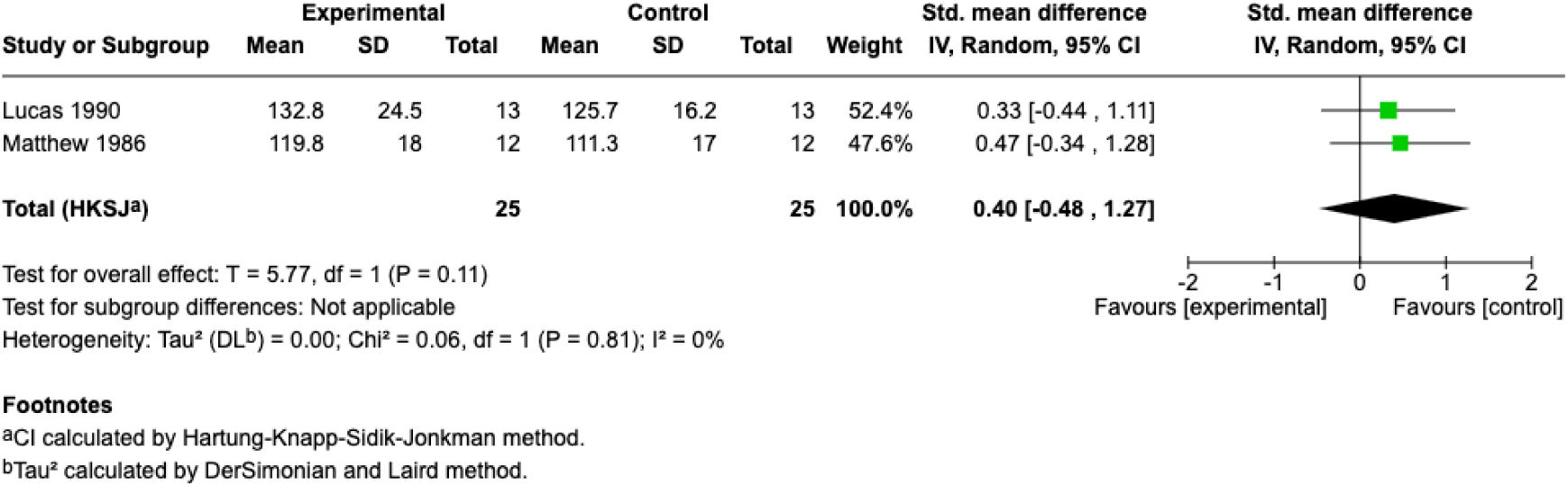
Forest plot of SBP comparing experimental vs control groups in two studies.

**Figure 7B:**
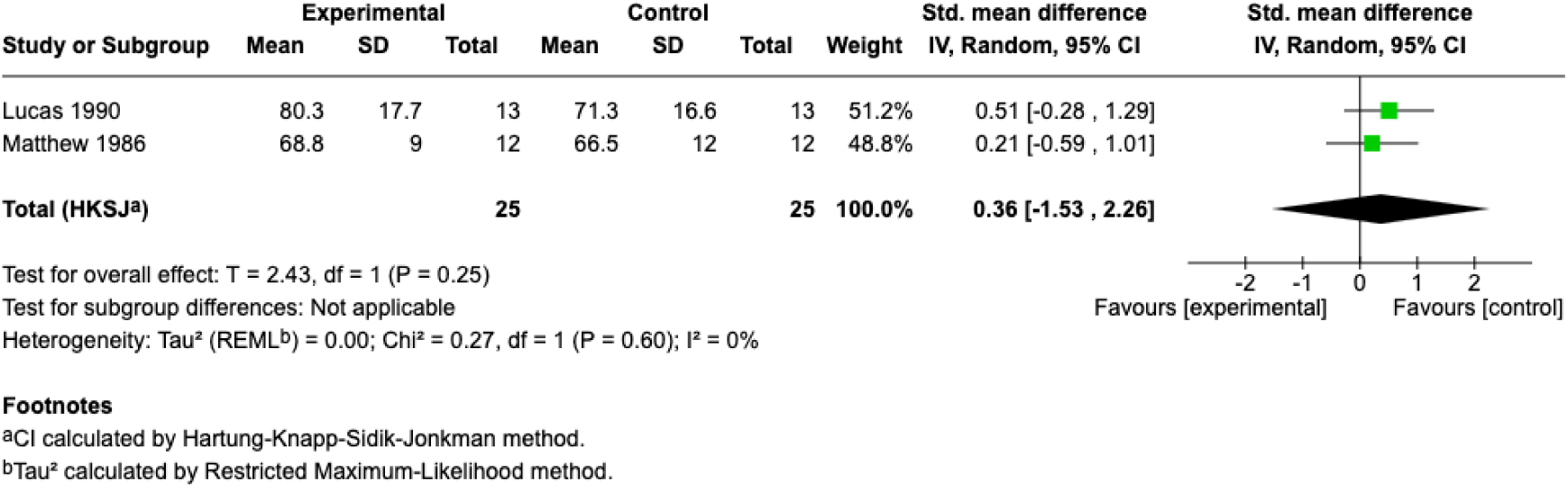
Forest plot of Pulse rate comparing experimental vs control groups in two studies.

**Figure 7C:**
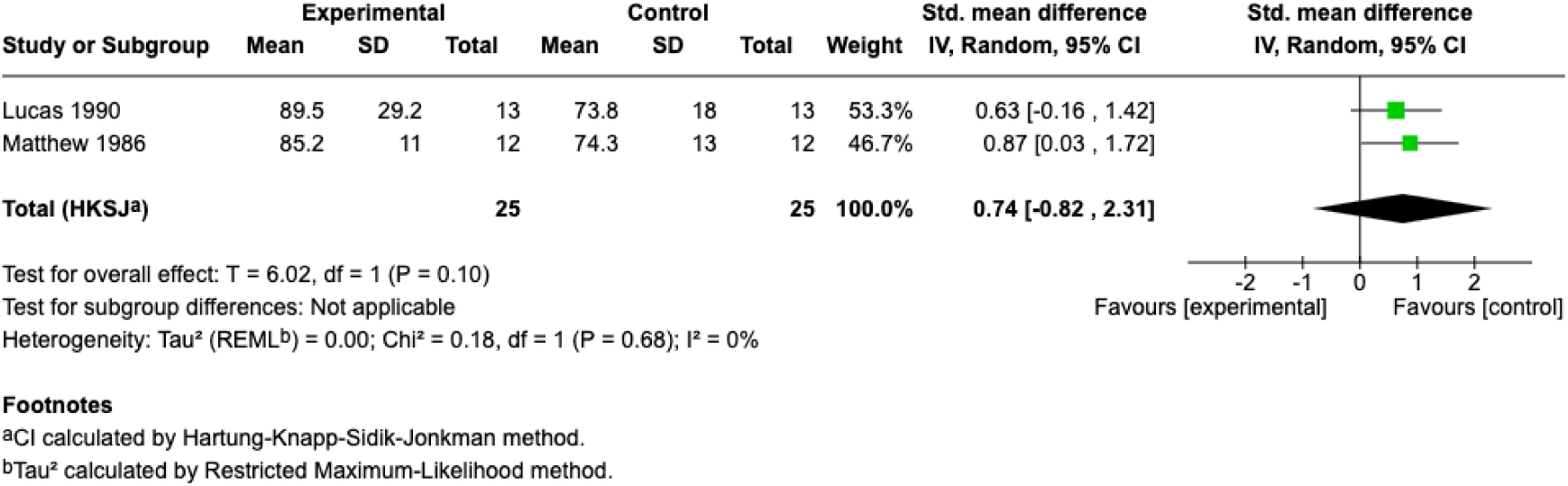
Forest plot of DBP comparing experimental vs control groups in two studies.

### 4.1 Discussion

This study was the first systematic review and meta-analysis to specifically explore the effects of caffeine on individuals with schizophrenia, examining its impact on cognitive function, symptom management, functional outcomes, and physiological measures. Overall, the evidence suggests that caffeine’s influence is complex and sometimes contradictory.

Caffeine’s impact on cognitive function was inconsistent. While some studies reported improvements in specific domains like working memory, motor speed, and visual memory, particularly among male patients (Núñez et al., 2015), other investigations found no significant benefit. These inconsistencies may reflect differences in sample characteristics, caffeine tolerance, baseline cognitive functioning, and methodological approaches. Prior studies in non-clinical populations suggest that caffeine’s cognitive-enhancing effects are often dose-dependent and influenced by individual sensitivity (Joshi, 2011, Ricupero and Ritter, 2024). In schizophrenia, where baseline neurocognitive deficits are common (Keefe and Harvey, 2012), caffeine may provide a modest benefit in specific domains but could also exacerbate attentional or executive dysfunction when overused.

Regarding symptom management, higher levels of caffeine intake were associated with fewer negative symptoms, but a trend toward increased positive symptoms. This is consistent with previous research suggesting that patients may use caffeine as a form of self-medication to counteract cognitive and motivational deficits (Williams and Gandhi, 2008, Wilkins, 1997). It also aligns with earlier research indicating that caffeine, as a stimulant, may exacerbate psychotic symptoms in vulnerable individuals (Cappelletti et al., 2015, Hotnauli and Husada, 2021, Henning et al., 2019). These dual effects may reflect caffeine’s dopaminergic action, alleviating negative symptoms through increased dopamine availability in prefrontal regions, but simultaneously worsening positive symptoms by enhancing dopaminergic activity in mesolimbic pathways (Broderick and Benjamín, 2004). However, our meta-analyses focusing on overall psychiatric symptoms, using measures like the BPRS and NOSIE, did not find significant differences between caffeine users and non-users.

Functional outcome findings were similarly variable, with some suggestion that limiting caffeine access in hospital settings is associated with improved global functioning and shorter hospital stays (Lagreula et al., 2023). Other studies did not confirm this relationship, arguing that higher caffeine consumption actually improved clinical function in standardised scales and made no difference in hospitalization outcomes (Mayo et al., 1993, Szoke et al., 2023).

Physiologically, caffeine consistently reduced cerebral blood flow—likely due to its adenosine receptor antagonism—but pooled analyses showed no significant effects on blood pressure or pulse rate. This contrasts with some studies in the general population linking caffeine to elevated cardiovascular measures, which may be explained by the chronic tolerance observed in many schizophrenia patients (Ringen et al., 2018, Mesas et al., 2011).

### 4.2 Clinical Implications

The findings of this review carry important clinical implications for the management of patients with schizophrenia. First, the observed association between caffeine consumption and reduced negative symptoms suggests that some patients may be self-medicating with caffeine. While this may offer short-term relief, clinicians should be mindful that caffeine use might also contribute to heightened positive symptoms, as well as potentially interfere with sleep and increase anxiety. Therefore, a balanced approach is necessary when advising patients on caffeine intake. The observation that restricting caffeine in inpatient settings was associated with better functional outcomes, including higher GAF scores at discharge and shorter hospital stays, suggests that limiting caffeine access in controlled environments could be beneficial for promoting recovery and reducing hospitalization time. This raises the possibility that clinical guidelines in psychiatric settings may need to consider moderating caffeine availability, especially among individuals with high daily intake or significant caffeine dependence. However, it is equally important to approach such restrictions cautiously and to provide supportive strategies to manage potential withdrawal effects, which could include irritability, headaches, or worsening of negative effects.

Moreover, since caffeine was found to significantly reduce cerebral blood flow and may impact brain physiology, clinicians should be cautious about high caffeine consumption, particularly in patients already vulnerable to neurovascular changes or those with comorbid cardiovascular risks. Although the meta-analysis on blood pressure and pulse did not reveal significant effects, individual variability in cardiovascular response suggests that personalized monitoring may still be necessary. Importantly, the lack of clear benefit on global psychiatric symptoms as measured by BPRS and NOSIE underlines that caffeine is unlikely to substitute or meaningfully augment antipsychotic treatment. Therefore, while it may be used by patients to cope with certain symptoms, healthcare providers should educate patients about its potential risks and interactions with antipsychotic medications, particularly as caffeine can influence the metabolism of drugs like clozapine and olanzapine.

### 4.3 Study Limitations

There were several limitations of this study. Firstly, many of the included studies had relatively small sample sizes, limiting the statistical power to detect more subtle associations and potentially inflating effect sizes. Substantial heterogeneity in the meta-analyses of psychiatric symptom outcomes suggests variability in study designs, populations, caffeine doses, and outcome measurements, making it difficult to draw definitive conclusions and complicating the generalizability of our findings.

Funnel plots were symmetrical but visual inspection may not be entirely conclusive. Moreover, the predominance of observational studies restricts our ability to infer causality. Residual confounders—such as variations in smoking behavior, concurrent antipsychotic medication use, and other substance-use patterns—were not uniformly controlled. Lastly, the scarcity of longitudinal data also precludes definitive conclusions regarding the long-term clinical and functional impact of caffeine consumption in this population.

### 4.4 Future Research

Future research should aim to address limitations in current literature with larger, well-powered randomised controlled trials. It would be particularly beneficial to standardize caffeine dosing and incorporate biochemical monitoring (e.g., serum caffeine levels) to clarify dose-response relationships. Further investigations should also explore the differential impact of caffeine based on symptom profiles, medication regimens, and comorbid conditions such as anxiety or cardiovascular risks. In addition, longitudinal studies could determine whether the short-term cognitive and symptomatic effects persist over time and to assess the potential impact on relapse rates. Finally, mechanistic studies examining how caffeine’s interactions with specific antipsychotic pharmacodynamics and neurobiological pathways would enhance understanding of the underlying processes driving these clinical observations.

## 5. Conclusion

This review highlights the complex and varied effects of caffeine on individuals with schizophrenia, particularly its potential to influence cognitive function, symptom management, and functional outcomes. While some evidence suggests beneficial effects on executive functioning and negative symptoms, other findings indicate a potential worsening of positive symptoms and minimal impact on physiological measures such as blood pressure and heart rate. These mixed results underscore the need for careful clinical consideration when managing caffeine consumption in this population. Given the current limitations in the evidence, including small sample sizes and study heterogeneity, further high-quality research is essential to clarify caffeine’s role and guide clinical recommendations for individuals with schizophrenia.

## Supporting information

Quality Assessment and Publication Bias

Supplementary table 1: Study Characteristics

## Data Availability

All data produced in the present study are available upon reasonable request to the authors

