## Supplementary material for "Caffeine Consumption and Schizophrenia: A Systematic Review and Meta-analysis of Cognitive, Symptomatic, and Functional Outcomes": Quality Assessment and Publication Bias

**Supplementary Tables 2 and 3**

**Table 2: Quality results for cohort studies**

| **Study** | **Item 1** | **Item 2** | **Item 3** | **Item 4** | **Item 5** | **Item 6** | **Item 7** | **Item 8** | **Item 9** | **Item 10** | **Item 11** | **Overall** |
| --- | --- | --- | --- | --- | --- | --- | --- | --- | --- | --- | --- | --- |
| Almis et al. (2023) | Yes | Yes | Yes | Yes | Yes | Yes | Yes | Yes | Yes | Yes | Yes | Good |
| Koczapski et al. (1989) | Yes | Yes | Yes | Yes | Yes | Yes | Yes | Yes | Yes | Yes | Yes | Good |
| Núñez et al. (2015) | Yes | Yes | Yes | Yes | Yes | Yes | Yes | Yes | Yes | Yes | Yes | Good |
| Szoke et al. (2023) | NA | NA | Yes | Yes | Yes | Yes | Yes | Yes | Yes | Yes | Yes | Good |

Item 1: Were the two groups similar and recruited from the same population?
Item 2: Were the exposures measured similarly to assign people to both exposed and unexposed groups?
Item 3: Was the exposure measured in a valid and reliable way?
Item 4: Were confounding factors identified?
Item 5: Were strategies to deal with confounding factors stated?
Item 6: Were the groups/participants free of the outcome at the start of the study (or at the moment of exposure)?
Item 7: Were the outcomes measured in a valid and reliable way?
Item 8: Was the follow up time reported and sufficient to be long enough for outcomes to occur?
Item 9: Was follow up complete, and if not, were the reasons to loss to follow up described and explored?
Item 10: Were strategies to address incomplete follow up utilized?
Item 11: Was appropriate statistical analysis used?

Answers: Yes/No/Unclear/NA

**Table 3: Quality results for cross-sectional studies**

| **Study** | **Item 1** | **Item 2** | **Item 3** | **Item 4** | **Item 5** | **Item 6** | **Item 7** | **Item 8** | **Overall** |
| --- | --- | --- | --- | --- | --- | --- | --- | --- | --- |
| Lagreula et al. (2023) | Yes | Yes | Yes | Yes | Yes | Yes | Yes | Yes | Good |
| Thompson et al. (2014) | Yes | Yes | Yes | Yes | No | No | Yes | NA | Fair |
| Topyurek et al. (2020) | Yes | Yes | Yes | Yes | Yes | Yes | Yes | Yes | Good |

Item 1: Were the criteria for inclusion in the sample clearly defined?
Item 2: Were the study subjects and the setting described in detail?
Item 3: Was the exposure measured in a valid and reliable way?
Item 4: Were objective, standard criteria used for measurement of the condition?
Item 5: Were confounding factors identified?
Item 6: Were strategies to deal with confounding factors stated?
Item 7: Were the outcomes measured in a valid and reliable way?
Item 8: Was appropriate statistical analysis used?

Answers: Yes/No/Unclear/NA
