## Supplementary table 1: Study Characteristics for "Caffeine Consumption and Schizophrenia: A Systematic Review and Meta-analysis of Cognitive, Symptomatic, and Functional Outcomes"

| **First author’s name and year of publication** | **Study design** | **Study region** | **Sample Size (% male)** | **Mean age (mean ± SD) years** | **Caffeine Exposure** | **Comparison Group** | **Outcomes measures** | **Results** |
| --- | --- | --- | --- | --- | --- | --- | --- | --- |
| Almis et al. 2023 | Cohort | Turkey | 177 (66.67) | 37.9 ± 9.95 | 454.3 ± 360.1 mg daily | 89 HC | SAPS, SANS | There was no significant correlations between daily caffeine consumption and SAPS or SANS scores in patients with schizophrenia. |
| Bissonnette et al. 2021 | RCT | Canada | 27 (74) | 25.3 ± 4.1 | 200 mg | 13 HC | PANSS, BNSS, PSYRATS | Results showed caffeine had no effect on alpha asymmetry in the SZ group, although caffeine produced a more global effect on the reduction of alpha2 power in the SZ group. Further, those with more positive symptoms were found to have a greater reduction in alpha2 power following caffeine administration. |
| Koczapski et al. 1989 | Cohort | Canada | 33 | NR | 18 highest caffeine consumers, averaging 14.4 cups daily, and the 15 lowest caffeine consumers, averaging 4.6 cups daily. | 15 lowest caffeine consumers | BPRS, NOSIE | No significant improvements in patients' behaviour occurred when decaffeinated coffee was first introduced, nor was there any deterioration in patients' behaviour when the regular coffee was reinstated. Only after decaffeinated coffee was reintroduced for the second time did any of the predicted changes in patients' behaviour occur. |
| Lagreula et al. 2023 | Cross-sectional | Belgium | Two different periods, the first (n = 142) (6.3) and the second (n = 119) (10.9) | 43 ± 12 | NR | 119 restriction of access to caffeine | GAF | After adjusting for potential confounders, reduced caffeine availability inside the hospital was significantly associated with higher Global Assessment of Functioning scores at discharge (adjusted odds ratio [aOR] = 2.86, 95% confidence interval [CI] = 1.77-4.62). |
| Lucas et al. 1990 | RCT | United States | 13 (92.3) | NR | 10 mg/kg | Each patient served as his/her own control; the order of placebo and active drug administration was determined by a randomization procedure. | BPRS | The change in BPRS total score was significantly higher after caffeine challenge than after placebo. Similarly significant increases were obtained for the BPRS subscales thought disorder and euphoria-activation. |
| Mathew et al. 1986 | RCT | United States | 24 (41.7) | 37.4 ± 14.5 | 250 mg | 12 placebo | Cerebral blood flow measurements and mental status examination | Caffeine produced significant cerebral blood flow reductions but no changes in the patient's clinical condition. Administration of caffeine was not accompanied by any changes in the patient's mental status. |
| Mayo et al. 1993 | RCT | United Kingdom | 26 (61.5) | 51 ± 18 | 160-1200 mg per cup | Placebo | MADRS, BPRS, NOSIE, CAS | No correlation was found between caffeine consumption and levels of anxiety and depression. No significant changes in patients' behaviour or levels of anxiety and depression occurred when the wards changed to decaffeinated products. |
| Núñez et al. 2015 | Cohort | Spain | 113 (61.9) | 45.63 ± 10.03 | NR | 61 HC | SAPS, SANS Verbal fluency, processing speed, and working, visual and verbal memory | Caffeine intake had beneficial effects on male schizophrenic patients only in complex tasks requiring deeper cognitive processing (semantic fluency, cognitive speed, working memory, and visual memory). Female patients and controls were unaffected. |
| Szoke et al. 2023 | Cohort | France | 804 (73.9) | 31.3 ± 9.2 | 2.2 cups/day | NA | PANSS, GAF, CGI | After controlling for potential confounders (demographic variables, smoking) only the negative dimension of psychosis was associated with the amount of caffeine ingested. Less severe negative symptoms were associated with higher caffeine consumption. The effect size of this association was small (partial correlation coefficient = -0.12) but significant. |
| Thompson et al. 2014 | Cross-sectional | Australia | 20 (85) | 42 | 523 mg per day | NA | NR | Participants’ behaviours related to caffeine consumption seemed to be tempered by their previous experiences of consumption; if participants had experienced positive effects such as alertness or relaxation in the past, their use was maintained at a similar level or increased. Conversely, participants who anticipated negative consequences often altered their patterns of caffeine consumption; for example, by substituting caffeinated drinks that minimised or ceased their experience of negative side effects for those that directly caused such impacts. |
| Topyurek et al. 2020 | Cross-sectional | Canada | 27 (77.8) | 27.3 ± 7.8 | 526.2 mg/day | 13 moderate caffeine users (≤ 250 mg/day) | Processing speed, executive function, working memory, sustained attention, visual learning  and memory, and verbal learning and memory. CSB, PANSS | Moderate caffeine users, compared to high caffeine users, demonstrated better performance on a task measuring executive function. While high caffeine users had fewer negative symptoms, they had more positive symptoms than moderate caffeine users. |

**NOTE**: SAPS = scale for the assessment of positive symptoms, SANS =scale for the assessment of negative symptoms, mg= milligram, PSYRATS= Psychotic Symptom Rating Scale, BNSS = Brief Negative Symptom Scale, PANSS = Positive and Negative Symptom Scale, RCT = Random controlled trial, SZ = schizophrenia, HC = health controls, BPRS = Brief Psychiatric Rating Scale, NOSIE = Nurses Observation Scale for Inpatient Evaluation, NR =Not reported, CAS = Clinical Anxiety Scale, MADRS = Montgomery Asberg Depression Rating Scale, GAF = Global Assessment of Functioning scores, CGI = Clinical Global Impression, CSB = Cogstate Schizophrenia Battery
